# The evaluation of the insertion parameters and complications of the i-gel Plus airway device for maintaining patent airway during planned procedures under general anaesthesia: a protocol for a prospective multi-center cohort study

**DOI:** 10.1101/2021.07.19.21260747

**Authors:** Olga Klementova, Taranpreet Kaur Bhoday, Jakub Werner, Ana M. Lopez, Will Donaldson, Erik Lichnovsky, Tomasz Gaszynski, Tomas Henlin, Jan Bruthans, Jitka Ulrichova, Martin Lukes, Jan Blaha, Michal Kral, Lidia Gomez, Raquel Berge, Jonathan Holland, Francis McAleavey, Shiva Arava, Lubica Merjava Skripecka, Sebastian Sobczyk, Tomas Tyll, Pavel Michalek

## Abstract

**Introduction:** Supraglottic airway devices represent a less invasive method of airway management than tracheal intubation during general anaesthesia. Their continued development is focused mainly on improvements in the insertion success rate and minimalization of perioperative and postoperative complications. The i-gel Plus is a novel, anatomically preshaped supraglottic airway device which achieves a perilaryngeal seal due to a non-inflatable cuff made of a soft thermoplastic elastomer. The purpose of this trial is to assess the success rate of the i-gel Plus use during elective procedures under general anaesthesia, its intraoperative performance, and the degree of postoperative complications.

**Methods and analysis:** This is a multicenter, prospective, interventional cohort study. The enrolment will take place in seven centers in four European countries. We plan to enrol 2000 adult patients in total, who are scheduled for elective surgery under general anaesthesia, and with an indication for use of a supraglottic airway device for management of their airway. The study is projected to run over a period of 18 months. The primary outcome of the study is the total success rate of the i-gel Plus insertion in terms of successful ventilation and oxygenation through the device. Secondary outcomes include perioperative parameters, such as insertion time, seal/leak pressures, number of insertion attempts, and postoperative adverse events and complications. Postoperative follow-up will be performed at 1 hour, 24 hours in all patients, and for selected patients at 3 and 6 months.

**Ethics and dissemination:** The trial has received the following ethical approvals: General University Hospital Prague, University Hospital Olomouc, University Military Hospital Prague, University Hospital Barcelona, University Hospital Lodz, Antrim Area Hospital, Craigavon Area Hospital, Office for Research Ethics Committees Northern Ireland. The results will be published in peer-reviewed journals and presented at relevant anaesthesia conferences.

**Trial registration number:** ISRCTN86233693

**ARTICLE SUMMARY:** *Strengths and limitations of this study:*

- The study will provide the first data about the use of the novel supraglottic airway device, the i-gel Plus for maintaining ventilation and oxygenation during anaesthesia.
- Broad inclusion criteria and the multicenter, multinational design of the study will allow study of a wide spectrum of populations of different age, gender and ethnicity and thus provide more generalisable results.
- Patients showing significant postoperative adverse effects will be followed up for up to 6 months which will provide information about the long-term complication rates of the device.
- The main limitation of the study is that the device will be trialled only in selected elective procedures and not as a rescue device or in difficult airway management scenarios.

## INTRODUCTION

Supraglottic airway devices are an integral part of airway management during general anaesthesia,^1^ in fact over half of all procedures under general anaesthesia are managed with a supraglottic airway in the UK.^2^ As a less invasive alternative to tracheal intubation,^3^ supraglottic devices are commonly used in general anaesthesia of elective operations and are increasing in popularity across the world. The first supraglottic device, the laryngeal mask airway, was developed in the 1980s by Dr. Archibald Jeremy Brain^4^ and was comprised of an elliptical inflatable cuff attached to an airway tube. His original design paved the way for a series of alterations and new inventions in the following years.^5-9^

There are many types of supraglottic airway devices available on the market today. They can be broadly classified into base of tongue sealers and perilaryngeal sealers,^10^ with or without an inflatable cuff.^11^ Supraglottic airway devices also differ in the mechanismu sed to achieve this seal - they may rely on cuff inflation, a wedge seal, or can be self energising.^12^ These devices can also be divided according to Tim Cook’s classification^13^ into first and second generation devices – second-generation devices possess an additional drainage channel for gastric content whereas first generation airways do not. It has been proposed that second-generation devices may be more protective against aspiration of gastric contents.^14^

The original i-gel supraglottic airway device was developed in 2007 by Dr. Nasir^15^ and was designed as a single-use, latex-free, second-generation supraglottic airway device with an innovatively shaped, non-inflatable cuff.^16^ This cuff anatomically mirrors the perilaryngeal anatomy and is made of a soft, gel-like thermoplastic elastomer, which apposes the perilarygneal anatomy to create a seal, therefore mitigating the need for inflation of a cuff and thus reducing pressure trauma to the local mucosa. Furthermore, by removing the requirement to inflate a cuff, the i-gel is easier and faster to insert than many devices and has better post insertion stability as there is no movement that may accompany the inflation of the cuff. The i-gel also includes an epiglottic rest to prevent epiglottic downfolding, an additional gastric channel for passive drainage of the gastric contents or insertion of the gastric tube, and a firmer stem with a bite block and curved buccal stabiliser leading to a multifunctional 15mm connector.

Initially designed for the airway maintenance during anaesthesia in elective procedures,^17^ the indications for the i-gel use have extended over time to include use in cardiopulmonary resuscitation^18^ or as a conduit for fiberscope-guided tracheal intubation.^19^

Several systematic reviews and meta-analyses have been published on the i-gel supraglottic airway device. The device exhibited similar oropharyngeal leak pressures, insertion time, and insertion success on the first attempt as the LMA Supreme™, while the LMA Supreme™ was associated with easier gastric tube insertion but more sore throat postoperatively.^20^ When systematically compared with all laryngeal mask airways, the i-gel has showed significantly less reports of postoperative sore throat and better fibreoptic views through the device whereas other parameters were without difference.^21^ Systematic reviews and meta-analyses comparing the i-gel with the LMA ProSeal™ did not find any differences in insertion parameters, apart from shorter time for insertion of the i-gel, higher seal pressures for the LMA ProSeal™, but significantly lower postoperative complaints in the i-gel patients.^22,23^

The i-gel Plus represents the second generation of the i-gel device. It is a novel cuffless, CE marked, anatomically shaped perilaryngeal wedge sealer with an additional wide gastric channel, allowing for the insertion of a gastric tube when required.

### Current knowledge

The i-gel® Plus from Intersurgical is a new 2^nd^ generation supraglottic airway with a non-inflatable cuff. When compared to a standard i-gel®, the device incorporates a number of additional features to enhance product performance. This includes an increase in the diameter of the gastric channel to allow for the insertion of a larger gastric tube. Ramps located at the end of the airway channel where it exits into the bowl of the non-inflatable cuff to optimise performance as a conduit for intubation, and a small increase in the length of the cuff tip to increase the oesophageal seal.

The device also includes features already incorporated into the i-gel O2 resus airway. A supplementary oxygen port for the delivery of passive oxygenation as part of a cardio-cerebral resuscitation (CCR) protocol and a hook ring for use with a specially designed airway support strap (available separately) to provide an alternative method for securing the device.

### Study aims

The primary objective for this study is to assess the total success rate of i-gel Plus insertion within a maximum of 3 attempts in adult patients indicated for elective surgical procedures without the need for muscle relaxation. Success is defined by effective oxygenation (defined as over 92% on pulse oximetry) and ventilation without a significant (audible) leak.

The study aims to meet several secondary objectives including assessing the time of insertion, seal/leak pressure obtained, and peri/post operative complications of the i-gel Plus.

## METHODS AND ANALYSIS

### Study protocol

The study protocol (version 1.2, 1/10/2019) has been created in concordance with the Declaration of Helsinki. Methodologically, it has been prepared using the following guidelines: SPIRIT 2013 Statement,^24^ SPIROS 2020 Recommendation,^25^ and STROBE Statement.^26,27^ This study is a prospective cohort, interventional, multicentre study.

The study is conducted in 7 hospitals (5 university and 2 general) in 4 countries (Czech Republic, Spain, Poland, and the United Kingdom). Anaesthesiologists will indicate their experience with the original i-gel device on the CRF. 4 groups have been created to differentiate between the more and less experienced operators: 0 previous insertions of the original i-gel, between 1 and 20, 21 to 50 and more than 51. It is expected that many anaesthesiologists will have some experience with using the original i-gel device, however, inexperienced clinicians are able to join the study as well. All operators will receive The i-gel Plus user guide in advance, will have an opportunity to watch the video with the i-gel Plus insertion and practice insertion on a manikin. Patients will be pre-selected from the surgical list at pre-anaesthetic clinic or a day before the scheduled operation and will be followed up 24 hours post-op in person. Long-term complications will also be searched for at 3 and 6 months after the operation by telephone to follow up persistence and improvement of symptoms.

### Eligibility criteria

Inclusion criteria: Patients requiring general anaesthesia with oxygenation and ventilation for elective surgeries that primarily do not require muscle relaxation and who can be managed with a supraglottic airway device will be recruited. Pregnant patients may also be included provided there are no other contraindications to management with a supraglottic device and they do not fulfill any of the exclusion criteria.

#### Exclusion criteria

➢ Age <18 or >89
➢ Acute procedures/non-fasted patients
➢ Increased risk for aspiration of gastric contents
➢ ASA > III
➢ BMI > 35kg/m^2^
➢ Unusual operation position (steep head down, prone, sitting)
➢ Unable to provide informed written consent
➢ Shared airway procedures (ENT, maxillofacial surgery, bronchoscopy)
➢ Intracavitary surgeries (laparotomies, thoracotomies, neurosurgery)

### Interventions

The study flow chart is shown in Figure 1. Eligible patients will undergo induction of general anaesthesia and the i-gel Plus device will be used to maintain a patent airway for oxygenation and ventilation throughout the procedure. Once the i-gel Plus is inserted its position in relation to the vocal cords will be evaluated with fiberoptic visualisation if adequate equipment is available. The gastric tube will be inserted through the gastric channel of the device if indicated (stomach distension, laparoscopic surgery, extended procedures).

**Figure 1.**
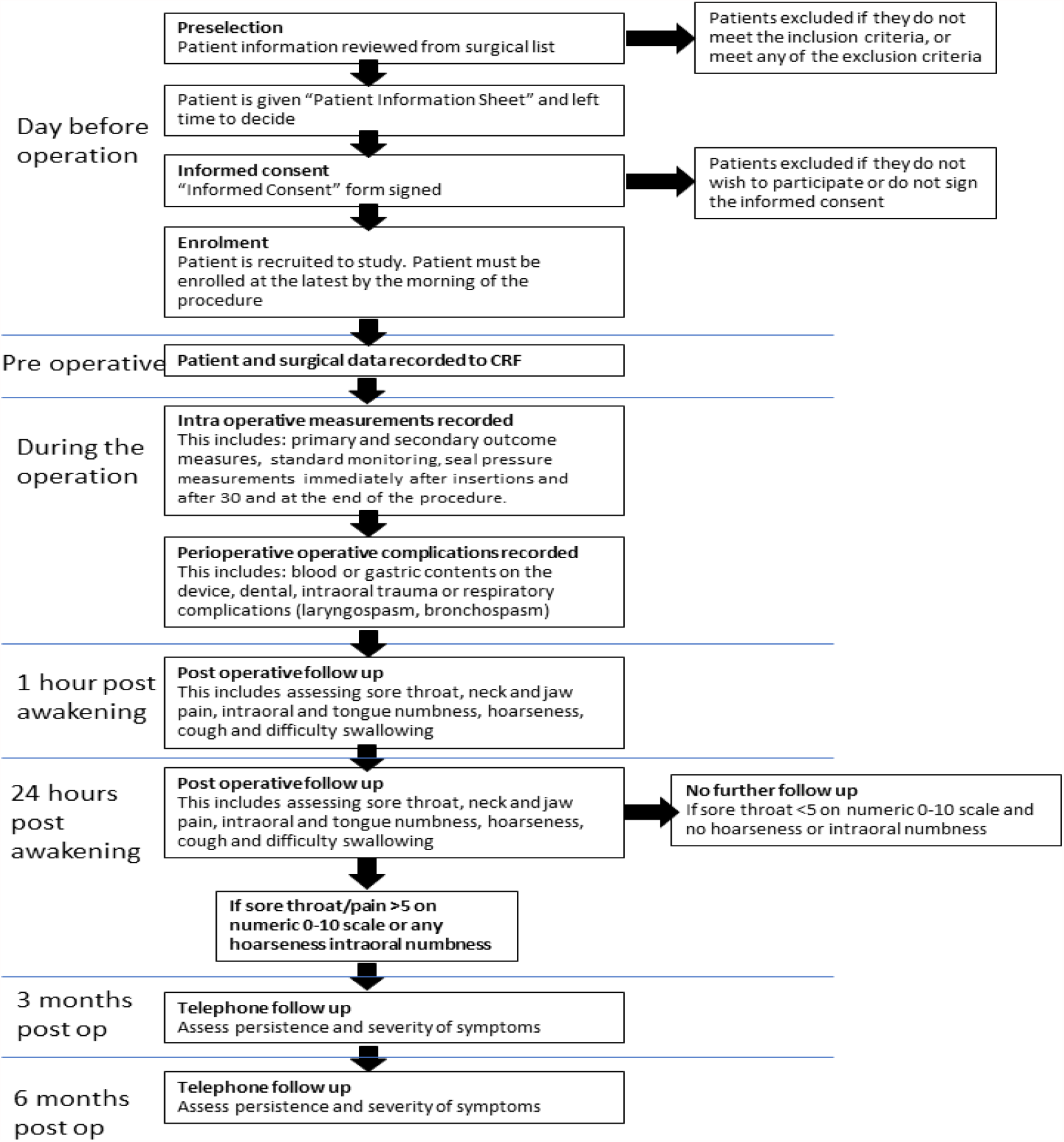
The study flow chart.

### Concomitant care

All patients will undergo intravenous induction of general anaesthesia using propofol and an opioid analgesic. Any other drugs administered will be recorded on the case report form. Patients are then monitored intraoperatively – using noninvasive blood pressure monitoring, ECG, pulse oximetry, and capnography. Invasive pressures will be monitored where indicated.

### Outcome measures

Primary outcome measure: The primary outcome measure is success of device insertion (within 3 attempts). An attempt is defined as the i-gel Plus passing through the teeth (or through the gums in toothless patients). If the device insertion is unsuccessful in 3 attempts, operators may choose to change the size of the i-gel Plus, in which case insertion may be attempted a further 3 times and recorded as successful, if it is. Alternatively, operators can choose to use a different supraglottic device or tracheal intubation, both of which would be recorded as failures for this study.

#### Secondary outcome measure

##### Intraoperative outcome measures

➢ Number of insertion attempts (including changing size)
➢ Insertion time (from moment tip crosses teeth until the device is connected to the anaesthetic machine)
➢ Time without oxygenation (from moment face mask ventilation is terminated until the first capnograph curve is visualised)
➢ Lowest saturation of haemoglobin during induction of anaesthesia and device insertion (measured by a pulse oximeter)
➢ Seal pressure (pressure at which leak is audible with stethoscope at the jugular area of the neck up to 40cm H_2_O on the Adjustable Pressure Limiting valve. If no leak is audible, the pressure is recorded as 40cm H_2_O)
➢ The subjective difficulty of insertion (using a Likert scale from 1-very easy to 5 – very difficult, as described in the patient case form)
➢ Assessment of device position (using a flexible, fiberoptic bronchoscope inserted to the end of the device breathing channel). The scores will be: 1-full view of vocal cords, 2-partial view of vocal cords including arytenoids, 3-only epiglottis visible, 4-no airway structures visible.^28^
➢ Number and ease of insertion attempts of a gastric tube (if indicated). The subjective difficulty of insertion will be recorded on 1-5 Likert scale.

##### Postoperative outcome measure

➢ Presence of blood on the device immediately after removal
➢ Gastric content on a bowl of the device immediately after removal
➢ Clinical signs of aspiration, laryngospasm, and bronchospasm
➢ Postoperative complications including sore throat, difficulty swallowing, hoarseness of voice, changes in tongue/intra-oral sensitivity, external neck pain, jaw pain (all measured on a subjective scale 1-10), and presence and nature of cough, recorded at 1 hour and 24 hours after awakening.
➢ Patients that report a sore throat, pain on swallowing, neck pain, or jaw pain more than 5 on a 1 to 10 scale and any hoarseness of voice or change in tongue/intra-oral sensitivity at 24 hours are followed up via telephone at 3 and 6 months for presence/improvement of the aforementioned parameters

### Recruitment

Patient recruitment began on the 21st of September 2020. Patients are pre-selected from the elective surgical list and screened for the exclusion criteria in advance. They receive the Patient Information Sheet at pre-anaesthetic clinic or the day before the surgery at admission. Patient recruitment is conducted by the study anaesthesiologist. Patients are included if they require general anaesthesia with oxygenation and ventilation for indicated elective surgeries that do not primarily require muscle relaxation.

### Data collection

Each hospital will be responsible for identifying patients potentially eligible for study recruitment. The study data are recorded onto a paper-based CRF by the anaesthesiologist or study nurse. Prior to the induction to anaesthesia, the patient data is recorded onto the paper CRF by the study nurse. All outcome measurements are recorded during and after the procedure by the study nurse. Any deviations from protocol or changes in surgery are recorded into the CRF including any adverse events observed, in which case the ethics committee will be informed. Every patient is allocated a unique study number which is used to pseudo-anonymise the data from the patient CRF and correlate it to an elecronic CRF which will be stored on a central RedCap database. The data management of the patients is in concordance with the national GDPR principles and is in adherence with the Declaration of Helsinki.

### Measurement of outcomes

The primary outcome measure is the total percentage success rate of device insertion. This will be calculated from categorical YES/NO success data from the CRF. The secondary outcomes will be measured and recorded according to the data types in Table 1.

**Table 1.** Secondary outcomes and their presentation

| Outcome | Type of Data |
| --- | --- |
| Number of insertion attempts | Numerical (1-3) |
| Insertion time | Continuous numerical (seconds) |
| Time without oxygenation | Continuous numerical (seconds) |
| Lowest saturation of hemoglobin | Continuous numerical (% SpO<sub>2</sub>) |
| Oropharyngeal seal pressure | Continuous numerical (cmH<sub>2</sub>O) |
| Subjective assessment of insertion difficulty | Numerical (1-5, Likert scale) |
| Fiberoptic assessment (if given) | Numerical (1-4 Score) |
| Insertion attempts of gastric tube (if required) | Numerical (1-5, Likert scale) |
| Insertion ease of gastric tube (if required) | Numerical (1-5, Likert scale) |
| Blood on device | Categorical (YES/NO) |
| Gastric content inside bowel | Categorical (YES/NO) |
| Clinical sign of aspiration | Categorical (YES/NO) |
| Laryngospasm | Categorical (YES/NO) |
| Bronchospasm | Categorical (YES/NO) |
| Sore throat | Numerical (0-10) and categorical YES/NO |
| Pain/difficulty on swallowing | Numerical (0-10) and categorical YES/NO |
| Hoarseness | Numerical (0-10) and categorical YES/NO |
| Intraoral/tongue numbness | Numerical (0-10) and categorical YES/NO |
| Neck pain | Numerical (0-10) and categorical YES/NO |
| Jaw pain | Numerical (0-10) and categorical YES/NO |

### Sample size calculation

The required sample size was calculated according to the modified Cochran formula^29^ assuming a minimum 95% success rate (in line with previously published 2^nd^ generation supraglottic airways device studies)^15,17,21,30-32^ and a 95% confidence level with a total confidence interval of 2%. This yielded a requirement for 1924 participants, and in order to compensate for potential participant dropout, we plan to collect at least 2000 patients.

Cochran formula

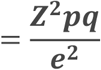

### Statistical analysis plan

For the statistical analysis, the SPSS statistical software, version 22 (SPSS Inc., Chicago, Illinois, USA) will be used. Data will be extracted from the electronic CRF by an independent statistician. Results are presented as a mean at 95% confidence level or a percentage.

A Fisher’s exact test will be used to analyse contingency tables of primary outcome data. Comparisons will be made between 3 operator groups defined by training stage (junior trainee with under 2 years practice, senior trainee with over 2 years practice, and staff grade/consultant) as well as for groups defined by experience with previous i-gel insertions (0; 1-20; 21-50; >50).

All numerical data including patient demographic data (gender, age, height, weight, BMI, and ASA classification) will be checked for normal distribution with the Shapiro-Wilk test and reported as either a percentage, median, or mean. These parameters along with modified Mallampati score, Simplified Airway Risk Index, mouth opening, neck circumference, limited neck movement, dentition status, and presence of beard will be presented as a mean or percentage and their effect on the performance of the i-gel Plus analysed using a manual logistic multivariable regression analysis. Each parameter will be individually eliminated if they do not contribute to the regression equation.

The secondary outcome data will be recorded as numerical (number of attempts, subjective assessment of insertion ease, fiberoptic assessment, and gastric tube insertion), continuous numerical (seal pressure and insertion time), or categorical (complications at the end of the procedure and postoperatively) data. Student t-test or Mann-Whitney U tests will be used for numerical data and Spearman’s rank correlation for any correlations.

### Methodological issues

The primary limitation of this study is that it does not cover the full spectrum of indications intended for the use of this novel device. The i-gel Plus is in this study trialled only in traditional indications of supraglottic airway devices and extended indications, such as insertion in shared airway surgery, prone positioning, resuscitation, emergency care, or difficult airway situations are not covered.

## ETHICS AND DISSEMINATION

The study received ethical approval from the Ethics Committee of the General University Hospital in Prague (study sponsor) in November 2019 (No. 1952/19 S-IV, received 14/11/2019, dr. J. Sedivy, Chair). The Czech Ministry of Health will act as a sponsor and partial founder via General University Hospital in Prague (grant number MZCZ-DRO-VFN64165). Following this initial approval, the ethical approvals from other centres were obtained: University Hospital Olomouc (No. 18/20, received 10/02/2020, Dr. J. Buresova, chair), University Hospital in Lodz (No. RNN/61/20/KE, received 03/03/2020, Prof. J. Drzewoski, chair), University Hospital in Barcelona (No. HCB 2020/0771, received 08/09/2020, Dr. A.L. Arellano Andrino, secretary), Northern HSC Trust for Antrim Area Hospital in Antrim (No. NT20-278410-10, received 10/12/2020, Dr. M. Rooney, Trust Director of Research and Development), Southern HSC Trust for Craigavon Area Hospital (No. ST2021/31, received 22/04/2021, Dr. P. Sharpe, Trust Director of Research and Development), Office for Research and Ethical Committee Northern Ireland (ORECNI) (REC 20/NI/0140, received 27/11/2020, Prof. P. Murphy, HSC REC B Chair), University Military Hospital in Prague (No. 108/15-104/2020, received 21/12/2020, Prof. J. Plas, chair). The study will be performed in compliance with the principles of the Declaration of Helsinki.

The study has been registered to the ISRCTN under number ISRCTN86233693 in November 2019 and protocol number U1111-1244-3085. Recruitment began in September 2020. This paper presents the initial protocol. Any potential modifications or ammendments of protocol must be approved by the Ethical Committees of the involved institutions.

### Data management, oversight, storage, and security

Designated main investigators at each site take a responsibility for the conduct of the trial. The study does not have a Data Monitoring Committee because no serious adverse effects are not expected and this was not requested by the Ethical Committee.

The completed paper CRFs will be secured in locked cabinets at each study site for 10 years. The data is pseudo anonymised using a unique study number and correlated to an electronic CRF stored on the RedCap database for review where it will be protected by a case-sensitive, alphanumeric password. After completion of the trial, the data will be shared using a public depository (https://data.mendeley.com).

### Dissemination plan

Preliminary (first 300 patients) and final study data will be presented at regional and international anaesthetic conferences, study website, mass media, and in scientific journals.

## Supporting information

SPIRIT checklist

## Data Availability

After completion of the trial, the data will be shared using a public depository (https://data.mendeley.com).

## Author contributions

PM, OK, WD, EL and TG conceived the study. TKB, JW, AML, TH, JB, JU, JB, JH, TT and PM designed and wrote the study protocol. ML, MK, LG, RB, FMA, SA, LMS and SS provided input into the final version of study protocol. OK, TKB, WD and PM drafted the manuscript. All authors read and approved the final version of the manuscript.

## Acknowledgments

The authors thank to the Intersurgical Ltd., David Chapman and Pete Seeney for providing detailed technical information on the device and for provision of the i-gel Plus devices for the study. The authors also thank Robert Greif and Lorenz Theiler for their input into the conceiving of the study.

## Patient and public involvement statement

It was not appropriate to involve patients or the public in the design, or conduct, or reporting, or dissemination plans of our research.

## Patient consent for publication

Not required

## Appendix

Patient information leaflet, patient consent form in English.

